# Evaluation of two commercial and two non-commercial immunoassays for the detection of prior infection to SARS-CoV-2

**DOI:** 10.1101/2020.06.24.20139006

**Authors:** Eric J. Nilles, Elizabeth W. Karlson, Maia Norman, Tal Gilboa, Stephanie Fischinger, Caroline Atyeo, Guohai Zhou, Christopher L. Bennett, Nicole V. Tolan, Karina Oganezova, David R. Walt, Galit Alter, Daimon P. Simmons, Peter Schur, Petr Jarolim, Lindsey R. Baden

## Abstract

**Background:** Seroepidemiology is an important tool to characterize the epidemiology and immunobiology of SARS-CoV-2 but many immunoassays have not been externally validated raising questions about reliability of study findings. To ensure meaningful data, particularly in a low seroprevalence population, assays need to be rigorously characterized with high specificity.

**Methods:** We evaluated two commercial (Roche Diagnostics and Epitope Diagnostics IgM/IgG) and two non-commercial (Simoa and Ragon/MGH IgG) immunoassays against 68 confirmed positive and 232 pre-pandemic negative controls. Sensitivity was stratified by time from symptom onset. The Simoa multiplex assay applied three pre-defined algorithm models to determine sample result.

**Results:** The Roche and Ragon/MGH IgG assays each registered 1/232 false positive, the primary Simoa model registered 2/232 false positives, and the Epitope registered 2/230 and 3/230 false positives for the IgG and IgM assays respectively. Sensitivity >21 days post symptom-onset was 100% for all assays except Epitope IgM, but lower and/or with greater variability between assays for samples collected 9-14 days (67-100%) and 15-21 days (69-100%) post-symptom onset. The Simoa and Epitope IgG assays demonstrated excellent sensitivity earlier in the disease course. The Roche and Ragon/MGH assays were less sensitive during early disease, particularly among immunosuppressed individuals.

**Conclusions:** The Epitope IgG demonstrated good sensitivity and specificity. The Roche and Ragon/MGH IgG assays registered rare false positives with lower early sensitivity. The Simoa assay primary model had excellent sensitivity and few false positives.

**Summary:** SARS-CoV-2 immunoassays can be valuable tools for informing the global response, but many currently available assays have not been independently validated. We conducted a performance assessment of four assays including the Roche Diagnostics and Epitope Diagnostics immunoassays.

## Background

In response to the rapid global spread of a novel virus, severe acute respiratory syndrome 2 (SARS-CoV-2), many commercial and research assays have been developed for the detection of acute or prior infection with SARS-CoV-2. Serological assays to detect antibodies to SARS-CoV-2 have received attention due to the large number of immunoassays being used for a range of purposes despite suboptimal validation [1–3].

Immunoassays are powerful tools that can be used to characterize the epidemiology and immunobiology of multiple pathogens including SARS-CoV-2 [4–8], identify suitable convalescent plasma donors [9], and potentially provide additional approaches for the clinical diagnosis of acute infection [10]. Seroepidemiological uses of these tools are numerous and include characterizing population-level seroprevalence [6]; identifying the fraction of asymptomatic and unreported infections [11]; informing group-specific and population-level control interventions [12]; monitoring transmission dynamics and the impact of control interventions over time [6,13]; and informing core epidemiological parameters such as the basic and effective reproductive numbers and infection and case fatality rates [14–17]. Serology-based cohort studies that combine classical seroepidemiological approaches with deep immune profiling can characterize the nature and kinetics of the humoral response and inform key questions including risks for reinfection and the parameters of protective titers [1,8,10,18].

However, despite enormous potential to guide the global COVID-19 response, confidence in serological tests and consequently the results of seroepidemiological studies have been undermined by poor (or poorly defined) test characteristics [3]. Given the importance of vigorous and independent immunoassay cross validation, we report on the performance of two commercial and two non-commercial assays.

## Methods

### Ethical considerations

The use of study samples and data was approved by the Mass General Brigham (MGB) (previously Partners Healthcare System) Institutional Review Board.

### Study design

We conducted a head-to-head test performance study using two commercial and two non-commercial SARS-CoV-2 immunoassays where laboratories were blinded to sample group.

### Study samples

Two panels of positive and negative control samples were selected from the MGB Biobank, a biorepository that contains biological samples and linked demographic and clinical data from >117,000 patients enrolled through the MGB network [19].

#### Positive control panel

To assess sensitivity, we selected 68 positive control samples from 28 patients that had been hospitalized at the Brigham and Women’s Hospital (BWH) between March 30 and May 4, 2020, and that previously tested positive by SARS-CoV-2 reverse-transcriptase polymerase chain reaction (RT-PCR). Samples were collected a mean of 10.5 days (standard deviation 6.0 days) post-RT-PCR confirmation and 16.1 days (standard deviation 5.4 days) post-symptom onset (PSO). The median number of samples per individual was two (range 1-5) and the median interval between sample collection was three days (range 2-6 days). The median age of patients was 57 years (range 32-79) and 18/38 (47%) were female (Table 1).

**Table 1.** Demographics and medical history of negative and positive controls^1^.

| Measures | Negative controls<br>(n=232) | Positive controls |  |  |  |
| --- | --- | --- | --- | --- | --- |
|  |  | 8-14 days<br>(n=30) | 15-21 days<br>(n=29) | >21 days<br>(n=9) | All<br>(n=68) |
| Demographics |  |  |  |  |  |
| Age in year, median (range) | 55 (20 - 89) | 59 (37 - 79) | 56 (32 - 79) | 56 (32 - 78) | 57 (32 - 79) |
| Female sex, N (%) | 90 (39%) | 17 (57%) | 15 (52%) | 3 (33%) | 35 (51%) |
| Race, N (%) |  |  |  |  |  |
| White | 198 (85%) | 12 (40%) | 7 (24%) | 3 (33%) | 22 (32%) |
| Black | 9 (4%) | 7 (23%) | 12 (41%) | 3 (33%) | 22 (32%) |
| Asian or Pacific Islander | 5 (2%) | 2 (7%) | 2 (7%) | 2 (22%) | 6 (9%) |
| American Indian or Alaskan Native | 0 (0%) | 2 (7%) | 1 (3%) | 0 (0%) | 3 (4%) |
| Other or not recorded | 20 (9%) | 7 (23%) | 7 (24%) | 1 (11%) | 15 (22%) |
| Ethnicity, N (%) |  |  |  |  |  |
| Non-Hispanic | 212 (91%) | 23 (77%) | 22 (76%) | 8 (89%) | 53 (78%) |
| Hispanic | 10 (4%) | 4 (13%) | 4 (14%) | 1 (11%) | 9 (13%) |
| Other or not recorded | 10 (4%) | 3 (10%) | 3 (10%) | 0 (0%) | 6 (9%) |
| Level of care<sup>2</sup> |  |  |  |  |  |
| Intensive care | N/A | 14 (47%) | 18 (62%) | 8 (89%) | 40 (59%) |
| Past medical history, N (%) |  |  |  |  |  |
| Hypertension | 124 (53%) | 13 (43%) | 12 (41%) | 6 (67%) | 31 (46%) |
| Obesity | 74 (32%) | 14 (47%) | 11 (38%) | 4 (44%) | 29 (43%) |
| Coronary artery disease | 68 (29%) | 10 (33%) | 10 (34%) | 3 (33%) | 23 (34%) |
| Asthma | 58 (25%) | 7 (23%) | 8 (28%) | 5 (56%) | 20 (29%) |
| Malignancy | 56 (24%) | 8 (27%) | 4 (14%) | 0 (0%) | 12 (18%) |
| Diabetes mellitus | 51 (22%) | 6 (20%) | 9 (31%) | 4 (44%) | 19 (28%) |
| Liver disease | 37 (16%) | 3 (10%) | 3 (10%) | 2 (22%) | 8 (12%) |
| COPD | 36 (16%) | 0 (0%) | 0 (0%) | 0 (0%) | 0 (0%) |
| Transplant | 29 (12%) | 2 (7%) | 3 (10%) | 1 (11%) | 6 (9%) |
| Other immune compromised conditions | 13 (6%) | 0 (0%) | 2 (7%) | 1 (11%) | 3 (4%) |
| Cerebrovascular accident | 10 (4%) | 1 (3%) | 0 (0%) | 0 (0%) | 1 (1%) |

1. Each sample was considered as an independent data point for calculating the values in this table.
2. Samples were considered "Intensive care" if they were collected from patients that required ICU admission at any point during hospitalization. Only hospitalized patients provided positive control samples, therefore the non-ICU fraction is one minus the proportion of ICU.
COPD Chronic obstructive pulmonary disease

#### Negative control panel

To assess specificity, we selected 232 prepandemic negative control samples from the MGB Biobank collected between August 28, 2017 and September 26, 2019. The median age was 55 years (range 20-89) and 90/232 (39%) were female. To determine if recent respiratory infections may be associated with increased cross-reactivity and false positives, we selected negative controls with and without recent respiratory infections. Of the total 232 negative control samples, 100 were from individuals without recent respiratory illness; 31 from individuals with prior laboratory-confirmed respiratory infections; 101 from individuals with a recent clinical diagnosis of respiratory infections including upper respiratory tract infection (n=50) or viral (n=11), bacterial (n=20) or unspecified (n=20) pneumonia (Table 2) based on diagnoses recorded in the electronic health record between 1 and 31 days prior to sample collection.

**Table 2.** Clinical and confirmed respiratory viral infections among negative controls.

| Recent acute illness | Days prior to sample collection | Males | Females | Total |
| --- | --- | --- | --- | --- |
| None | N/A | 70 | 30 | 100 |
| Upper respiratory tract infection | 1-14 days | 15 | 10 | 25 |
|  | 15-31 days | 9 | 16 | 25 |
| Bacterial pneumonia | 1-14 days | 7 | 3 | 10 |
|  | 15-31 days | 5 | 5 | 10 |
| Viral pneumonia | 1-14 days | 3 | 2 | 5 |
|  | 15-31 days | 4 | 2 | 6 |
| Unspecified pneumonia | 1-14 days | 7 | 3 | 10 |
|  | 15-31 days | 7 | 3 | 10 |
| Confirmed viral respiratory infection<sup>1</sup> | N/A | 15 | 16 | 31 |
| **Total** |  | **142** | **90** | **232** |

N/A Not applicable or not available
<sup>1</sup> Includes Parainfluenza antigen positive (n=13), Metapneumovirus antigen (10), influenza A/B antigen (8), Influenza A PCR (3), Influenza B PCR (1), RSV antigen (n=5), RSV PCR (1), Adenovirus antigen (3), Herpes Simplex I (DFA). Total number add up to more than 31 as some individuals recorded >1 positive result.

To ensure valid comparison between assays and given differences in plasma/sera requirements according to manufacturer/assay specifications, we only selected controls with both serum and plasma available from the same individual and time point. All samples were stored at −80°C following sample processing and none underwent thaw-refreezing cycles prior to analysis. Except for sample type (i.e. serum or plasma), identical panels of positive and negative controls were provided to each of the four participating laboratories (with two fewer samples provided to one lab). Samples were blinded to all laboratory staff and were only unblinded after results were provided to the principal investigators.

### Clinical data

We searched an electronic health records system linked to the Biobank to extract demographic data for all controls and clinical data for positive controls, including chronic medical conditions, level of care (non-ICU versus ICU), and date of RT-PCR test. Two study investigators independently reviewed medical records to determine symptom onset date; symptoms included cough, fever, dyspnea, myalgias, new loss of taste or smell, or sore throat.

### Serological assays and protocols

We assessed the performance of four immunoassays including (i) Elecsys Anti-SARS-CoV-2 (Roche Diagnostics, Indianapolis, USA) immunoassay intended for the qualitative detection of antibodies against the nucleocapsid (NC) antigen [20] (thought to include IgG, IgM, and IgA, although IgM and IgA are not specified in product information); (ii) EDI New Coronavirus COVID-19 enzyme-linked immunosorbent assays (ELISA) (Epitope Diagnostics, USA) that detect IgG against the NC antigen [21] and IgM against an unspecified antigen[22]; (iii) Ragon/MGH in-house ELISA that detects IgG, IgM and IgA against the receptor binding domain (RBD); and (iv) the Single Molecule Array multiplex assay (Simoa) that detects IgG, IgM, and IgA against the spike protein, S1 subunit, RBD, and NC [10]. The Ragon/MGH assay was performed at the Ragon Institute of Massachusetts General Hospital, Massachusetts Institute of Technology and Harvard; all other assays were performed at the Brigham and Women’s Hospital. Commercial assays were performed according to manufacturer specifications. The Simoa and Ragon/MGH assays were performed according to previously described methods [10,23].

### Result classification

Threshold cutoffs for defining positive, negative or indeterminate/borderline test results were defined according to manufacturer specifications for commercial assays. Threshold cutoffs and result determination for the non-commercial assays were established by the respective laboratories prior to the study. The Ragon/MGH positive cutoff was equal to the mean of the OD-converted ug/ml values of the negative control wells on the respective plate plus three times the standard deviation of the concentration from over 100 pre-pandemic plasma samples. Background-corrected concentrations were divided by the cutoff to generate signal-to-cutoff (S/CO) ratios. Samples with S/CO values greater than 1.0 were considered positive. Given the Simoa multiplex assay includes 12 output measures per sample (IgG, IgM, and IgA against four viral epitopes), result determination was based upon three pre-study classification models trained using an independent panel of 142 positive samples by RT-PCR SARS-Cov-2 and 200 negative pre-pandemic controls. Two models used cross-validation to select the fewest number of informative markers. An “Early Model” encompassed all positive RT-PCR timepoints within one week following the positive RT-PCR in pre-pandemic controls. This cross-validation yielded four markers (IgA S1, IgA Nucleocapsid, IgG Nucleocapsid, and IgG Spike) and exhibited the best performance in the training set [10]. A “Late Model” encompassed all RT-PCR positive timepoints after one week since PCR against all pre-pandemic controls. This cross-validation yielded only a single marker IgA S1. The model-specific threshold for a positive test result was determined based on the cutoff that yielded 100% specificity in the training set. A third model was a simple “12 Parameter” logistic regression using all 12 outputs.

### Data analysis

We performed five primary independent analyses. One each for the Roche (which provides a single result for all isotypes) and Ragon/MGH (IgG) assays; two for the Epitope immunoassays (IgG and IgM); and one for the primary Simoa assay “Early Model”. Analyses of the Ragon IgA/IgM and Simoa “Late Model” and “12-Parameter Model” are included in supplementary materials. Indeterminate or borderline results were considered negative for all analyses. We determined sensitivity using positive controls and specificity using negative controls as the gold standard. Sensitivity was calculated independently for samples collected 8-14, 15-21, and >21 days from symptom onset. Assay agreement was calculated between the Roche, Ragon/MGH IgG, Epitope IgG, and Simoa Early Model using prevalence-adjusted and bias-adjusted Kappa [24]. Binomial exact 95% confidence intervals were calculated for all estimates. All analyses were performed using the R software package (Version 4.0, www.R-project.org/).

## Results

### Specificity

The Roche and Ragon/MGH (IgG) assays registered 1/232 false positive for specificities of 99.6% (95% CI 98.7-100%). The Epitope IgG and IgM assays registered 2/230 and 3/230 false positives for specificities of 99.1% (97.9-100%) and 98.6% (95% CI 97.2% - 100%) respectively. No Epitope false positives overlapped and therefore if combining the two assays to provide a single result, the specificity is lower (5/230 false positives; 97.8% [95% CI 95.9-100%]). The Simoa Early Model registered 2/232 false positives with a specificity of 99.1% (95% CI 97.9-100%) (Table 3). The Ragon/MGH IgM and IgA assays demonstrated lower specificity at 94% and 70% respectively; Simoa Late and 12-marker models demonstrated specificities of 98.7% and 94% (supplemental data). Of the combined nine false positives results (Roche [1], Ragon/MGH IgG [1], Epitope IgG [2] and IgM [3], and Simoa Early Model [2]), five were from 100 negative controls without recent respiratory infection and four from 132 negative controls with recent respiratory infection, suggesting cross reactivity due to recent respiratory infections is unlikely to be an important cause of false positives in these assays. No common coronaviruses (e.g. 229E, NL63, OC43 or HKU1) were documented among these samples, however, so these data do not assess for common coronavirus-specific cross reactivity. Sensitivities: All assays except the Epitope IgM demonstrated 100% sensitivity among samples collected >21 days PSO, although only nine samples were included in this time period (Table 4). For the remaining time periods, the Simoa Early Model registered the fewest false negatives and highest sensitivities. However, confidence intervals generally overlapped across assays included in the primary analyses except for the earlier time periods when the Simoa Early Model was more sensitive than the Roche, Ragon/MGH IgG and the Epitope IgM assays (Table 4). At 21 days PSO, the Ragon/MGH IgM and IgA assays demonstrated sensitivities of 77.8% and 100% respectively; Simoa Late and 12-paremeter models demonstrated 100% sensitivity (supplemental table). Among twelve Ragon/MGH IgG false negatives, 4/12 were from individuals that subsequently seroconverted; 6/12 were from three individuals that were receiving immunosuppressive therapy; and 2/12 were collected at 11 and 18 days PSO from patients without subsequent sample collections. Of seven Roche false negatives, 2/7 were from individuals that seroconverted in later samples; 3/7 samples were from two patients receiving immunosuppressive therapy; and the remaining 2/7 were from the same patient collected 16 and 18 days PSO. Of two Epitope IgG false negatives, one later seroconverted and one was immunosuppressed. Of 25 Epitope IgM false negatives, 5/25 later seroconverted, 6/25 were immunosuppressed, and 14/25 were collected 10-37 days PSO. The Simoa Early Model registered only one false negative that seroconverted in later samples. When excluding individuals receiving immunosuppressive therapy and early samples individuals that subsequently seroconverted, there were no false negatives for the Simoa assay or Epitope IgG assays, two false negatives apiece for the Ragon/MGH and Roche assays, and 14 false negatives for the Epitope IgM assay.

**Table 3.** Assay specificities.

| Immunoassay | No. of negative controls<sup>1</sup> | No. testing negative | % | 95% CI |
| --- | --- | --- | --- | --- |
| Epitope IgG | 230 | 228 | 99.1 | 97.9% - 100% |
| Epitope IgM | 230 | 227 | 98.7 | 97.2% - 100% |
| Ragon/MGH IgG<sup>2</sup> | 232 | 231 | 99.6 | 98.7% - 100% |
| Roche<sup>3</sup> | 232 | 231 | 99.6 | 98.7% - 100% |
| Simoa (Early)<sup>4</sup> | 232 | 230 | 99.1 | 97.9% - 100% |

1. Given limited negative control aliquots, the Epitope assays were tested against 230 samples, versus 232 for the remaining assays
2. For specificity of Ragon/MGH IgM and IgA see supplementary materials.
3. The Roche Elecsys Anti-SARS-CoV-2 immunoassay detects IgG and likely IgM and IgA; details of other isotypes are not provided by manufacturer
4. Specificity of the Simoa multiplex assay Early Model. For specificities of the Late and 12-Parameter Models see supplementary materials.

**Table 4.** Assay sensitivities by days post-symptom onset.

| Assay | Days PSO | No. of positive controls | No. testing positive | % | 95% CI |
| --- | --- | --- | --- | --- | --- |
| Epitope IgG | 8-14 days | 30 | 29 | 96.7 | 90.2% - 100% |
|  | 15-21 days | 29 | 27 | 93.1 | 83.9% - 100% |
|  | > 21 days | 9 | 9 | 100 | 100% - 100% |
| Epitope IgM | 8-14 days | 30 | 20 | 66.7 | 49.8% - 83.5% |
|  | 15-21 days | 29 | 20 | 69 | 52.1% - 85.8% |
|  | > 21 days | 9 | 3 | 33.3 | 2.5% - 64.1% |
| Ragon/MGH IgG<sup>1</sup> | 8-14 days | 30 | 23 | 76.7 | 61.5% - 91.8% |
|  | 15-21 days | 29 | 24 | 82.8 | 69.0% - 96.5% |
|  | > 21 days | 9 | 9 | 100 | 100% - 100% |
| Roche<sup>2</sup> | 8-14 days | 30 | 26 | 86.7 | 74.5% - 98.8% |
|  | 15-21 days | 29 | 26 | 89.7 | 78.6% - 100% |
|  | > 21 days | 9 | 9 | 100 | 100% - 100% |
| Simoa (Early)<sup>3</sup> | 8-14 days | 30 | 29 | 96.7 | 90.2% - 100% |
|  | 15-21 days | 29 | 29 | 100 | 100% - 100% |
|  | > 21 days | 9 | 9 | 100 | 100% - 100% |

1. For sensitivity of Ragon/MGH IgM and IgA see supplementary materials.
2. The Roche Elecsys Anti-SARS-CoV-2 immunoassay detects IgG and likely IgM and IgA; details of other isotypes are not provided by manufacturer
3. Sensitivity of the Simoa multiplex assay Early Model. For sensitivities of the Late and 12-Parameter Models see supplementary materials.

### Assay agreement

Inter-assay concordance for negative controls was high for all combinations of assays with the highest agreement between Roche and Ragon/MGH IgG assays (Kappa 0.98, 95% CI 0.94-1.00) and the lowest between Epitope IgG and Simoa Early Model (Kappa 0.97, 95% CI 0.91-0.99). Of nine total false positives across the five assays none were overlapping between assays. Inter-assay agreement for positive controls was more variable and ranged from Kappa 0.94 (95% CI 0.80-0.99) between the Simoa and Epitope IgG assays to 0.68 (95% CI 0.46-0.83) between the Ragon/MGH IgG and Epitope IgG and Roche assays. Lower concordance between positive controls was largely driven by the higher numbers of false negatives observed in the Ragon/MGH IgG and Roche assays. Of the 20 discrete false negative results, 10 overlapped between one or more assay.

## Discussion

We report on the test performance of two widely used commercial and two non-commercial SARS-CoV-2 immunoassays using a panel of 300 well characterized control samples including 68 positive control samples from 28 RT-PCR SARS-CoV-2 confirmed hospitalized patients and 232 pre-pandemic negative control samples that included 132 samples from individuals with recent respiratory infections. We assessed the sensitivity, specificity, and agreement between assays using pre-defined thresholds and methods. Because the sensitivity of serological assays is dependent on when samples are collected relative to symptom onset, we stratified assay sensitivity by time from symptom onset to sample collection. To our knowledge, this is the first peer-reviewed performance evaluation of the recently released Roche SARS-CoV-2 immunoassay and includes a comprehensive evaluation of the EDI Epitope assays. Unlike most head to head SARS-CoV-2 immunoasssay performance evaluations, unique patient-sample combinations were used for all assays.

All four assays performed well with small absolute differences between them. The Roche and Ragon/MGH immunoassays registered the lowest number of false positives (1/232) for a specificity of 99.6% (95% CI 98.7-100%). These findings align with the Roche assay package insert (10/5272 false positives, specificity 99.81% [95% CI 99.65-99.91%])[20] and confirm excellent specificity. The Simoa Early Model and Epitope IgM assay registered two false positives apiece with slightly lower specificity (99.1% [95% CI 97.9-100%]), although with overlapping confidence intervals; the Epitope IgM performed less well with lower specificity, as did other Simoa models and the Ragon/MGH IgM and IgA assays. The Epitope IgG assay specificity in this study was higher than other recent studies that reported specificities of 88.7% from 53 negative controls [25] and 89.8% from 108 negative controls [23]. The overall performance of Epitope is superior when considering only the single IgG rather than combining the IgG and IgM assays. Given that even small decreases in specificity can substantially decrease the positive predictive value (PPV), particularly in populations with low pre-test probabilities or with low true disease prevalence, our findings suggest the Epitope IgG assay alone is preferable, particularly if used for seroepidemiological purposes. For example, when testing a population with a 3% true prevalence of disease with a test that is 98% specific, the PPV will be approximately 60% and an estimated four in ten positive results will be false positives. Conversely, if using a test that is 99.5% specific applied to the same population, the PPV is approximately 85% and slightly over one in seven positive results will be false positives. Given that most populations globally are assumed to have low SARS-CoV-2 seroprevalence, likely <5% with substantial heterogeneity, small differences in test specificity are of critical importance [5,14].

With the exception of Epitope IgM, all assays demonstrated 100% sensitivity among samples collected >21 days PSO, although only nine samples were included from this time period. The sensitivity was lower with greater variability between assays for samples collected earlier after symptom onset. This is to be expected given serological assays for most pathogens including SARS-CoV-2 are well described as having poor sensitivity earlier in disease course. Current reported data suggests the mean interval from symptom-onset to seroconversion for SARS-CoV-2 is ∼13 days for both IgM and IgG, although substantial heterogeneity exists between individuals and assays; by 25-27 days PSO, 98-100% of cases are expected to be seropositive by most serological assays [26,27]. By comparison, the assays in this study demonstrated 67-100% sensitivity for samples collected between 8-14 days PSO and 69-100% sensitivity for samples collected 15-21 days PSO. The Simoa Early Model assay demonstrated exellent sensitivity during the earlier PSO windows. The sensitivity of the Roche assay was slightly higher during earlier periods and slightly lower later compared to data reported by the manufacturer; Roche reported 66%, 88% and 100% sensitivity at 0-6, 7-13 and ≥ 14 days post-PCR confirmation [20], versus our findings of 71%, 95%, and 94% using the same time and date-from PCR-confirmation criteria (data not shown). The sensitivity of the Epitope IgG assay was lower than package insert data that report 100% for 30 positive controls in the second week of disease [21] and lower than a German study that reported 100% sensitivity for 22 positive controls [25] but higher but with overlapping confidence intervals for one US-based study that reported sensitivities of 84% at 6-20 days and 91% at >20 days [23].

This study has several limitations. Although this study was based on well characterized controls, we cannot definitively extrapolate findings to other populations [28]. For example, the sensitivity of these assays was assessed in samples from RT-PCR confirmed hospitalized patients with samples collected a mean of 16 days (SD 5) after symptom onset. Given most SARS-CoV-2 infections are mild or asymptomatic and do not require hospitalization, and given that little is known about humoral kinetics >2 months post infection, the sensitivity of these assays may be lower in individuals > 2 months post-infection and/or individuals with mild or asymptomatic infections that likely mount less durable immune responses [26,28]. We did not include negative control samples from individuals with confirmed common coronaviruses, which may increase cross reactivity and false positives.

Our study findings align with manufacturer data for the Roche Diagnostics’ Elecsys Anti-SARS-CoV-2 IgG immunoassay and confirm that false positives are rare, specificity is high, and sensitivity is excellent > 3 weeks after symptom onset, but lower early in the disease course and among immunocompromised individuals. The Epitope Diagnostics IgG immunoassay has lower specificity and sensitivity than data released by the manufacturer, but overall performed well with few false positives and high sensitivity. The Epitope IgM had more false positives and lower sensitivity and given IgM does not appear to rise substantially earlier than IgG in SARS-CoV-2 infections [26], the Epitope IgM assay may not add value to the IgG assay alone. The Ragon/MGH IgG immunoassay largely aligned with the Roche assay and false positives were rare, specificity was high, and sensitivity was excellent more than three weeks post-infection but lower earlier in disease course and among immunosuppressed individuals. The Simoa Early Model demonstrated high specificity and excellent sensitivity.

## Conclusions

Characterizing the epidemiology and immunobiology is a key priority for informing and responding to the current SARS-CoV-2 pandemic. For this we need test performance studies such as this to reliably and independently validate immunoassays.

## Data Availability

The data that support the findings of this study are available from the corresponding author, upon reasonable request.

## Acknowledgements

This work was largely funded by Brigham Health. EN is supported by a CDC U01 GH002238. LB is supported by NIH UM1AI069412 and UL1TR001102. D.S. is supported by NIH K08 AR075850.

